# The quantification of daily carbohydrate periodization among endurance athletes during 12 weeks of self-selected training: presentation of a novel Carbohydrate Periodization Index

**DOI:** 10.1101/2022.06.21.22276725

**Authors:** Jeffrey A. Rothschild, James P. Morton, Tom Stewart, Andrew E. Kilding, Daniel J. Plews

## Abstract

**Background:** Contemporary sports nutrition guidelines recognize that endurance athletes should periodize their daily carbohydrate (CHO) intake according to the demands of their training and competitive schedule. However, objective assessments of the dietary CHO periodization practices adopted by endurance athletes during prolonged training periods are not readily available.

**Objectives:** To objectively assess the magnitude of the CHO periodization practices adopted by endurance athletes via the formulation of a novel CHO Periodization Index (CPI). The CPI is proposed to represent a single metric to quantify how tightly an athlete’s CHO intake is matched with training load, the magnitude of adjustment, and how frequently these adjustments occur.

**Methods:** Self-selected training and dietary intake was reported daily by 55 endurance athletes (61.8% male) for 12 weeks (representing a total of 4,395 days of dietary assessments). Calculations were made for correlations between daily CHO intake and training load (product of session rating of perceived exertion and duration), CHO monotony (mean daily CHO intake divided by SD), CHO range (highest minus lowest single-day intake), and the CPI (correlation * range / monotony). Sub-group analysis was also performed to examine differences in CPI, frequency of fasted training sessions, and weekly training volume based on competitive level, habitual diet, and sex.

**Results:** Mean participant daily CHO intake was 3.9 ± 1.5 (range 1.2 to 7.2) g/kg, with the highest single-day intake being 17.6 g/kg. Mean CHO range was 6.6 ± 3.1 (range 2.0 to 15.2) g/kg. Carbohydrate monotony values ranged from 1.0 to 6.0. Pearson correlations between training load and daily CHO intake ranged from −0.34 to 0.87. Mean CPI was 1.0 ± 1.2 (range - 1.2 to 5.6) and was higher among the highest-level athletes.

**Conclusion:** Endurance athletes do not readily adjust daily CHO intake according to the demands of training. Furthermore, the CPI represents a promising tool that that can be used by researchers, coaches, and athletes to quantify CHO periodization practices and compare within and between individuals.

**Key Points:**

- It is recommended that endurance athletes adjust their daily carbohydrate intake according to variations in exercise volume and intensity, but there is limited knowledge of how this is being applied by athletes, and limited methods for quantifying or assessing the variations in intake.
- We introduce a novel Carbohydrate Periodization Index (CPI), a single metric to capture how tightly an athlete’s carbohydrate intake is adjusted based on training load, the magnitude of adjustment, and how frequently these adjustments occur.
- Data demonstrate that many endurance athletes do not follow recommended practices of adjustment in daily carbohydrate intake, or if they do, the magnitude of adjustment is small relative to changes in training volume and/or intensity.

## 1. Introduction

A periodized carbohydrate diet is a dietary pattern in which carbohydrate intake is varied according to the type of session and its goals within a periodized training cycle [1]. This practice is in line with contemporary sports nutrition guidelines recommending carbohydrate intake be individualized to the athlete and their event, and modulated according to changes in exercise volume [2]. These alterations to daily carbohydrate intake have the potential to modulate cell signaling pathways that regulate training-induced skeletal muscle adaptations [3, 4], influence training intensity and exercise capacity [5–7], and reduce the risk of inadequate energy availability [8]. At its simplest level, carbohydrate periodization can be recognized practically by daily fluctuations in total carbohydrate intake that consider the physical demands of the upcoming training and competitive schedule.

Despite the strong theoretical rationale for periodizing dietary carbohydrate intake, there is limited evidence of how such practices are currently practiced by athletes during real-world, day-to-day training. To date, knowledge of athlete practices has largely been limited to surveys [9–12], case studies [13, 14], or short-duration (∼7 d) observations [15]. In a survey of nearly 2,000 endurance athletes across a range of competitive levels, less than half reported eating differently depending on the duration or intensity of the upcoming exercise session and only ∼10% reported following a periodized carbohydrate dietary pattern (defined as adjusting carbohydrate intake based on their planned training sessions) [12]. In surveys of elite runners and race walkers, 22% of athletes reported following a periodized carbohydrate dietary pattern [10], and although 63% reported eating more food/calories on hard training days, only 20% reported eating less on easy training days [9]. It has also been found that despite an awareness of dietary periodization strategies, they were not widely implemented by elite runners and race walkers during one week of intensified training [15]. This suggests there may be a gap between what athletes are doing and the current best-practice recommendations of periodized nutrition.

Although the concept of fueling for the work required is easy for an athlete to understand, the degree to which the diet and training are periodized is likely to vary greatly among athletes. Combined with the lack of precision in questionnaires (e.g., simply asking someone if they adjust carbohydrate intake based on their planned training sessions), these findings highlight a need to objectively quantify the degree of periodization occurring in the context of an athlete’s training load. Accordingly, the purpose of this observational study was to report self-selected dietary intake in endurance athletes across a 12-week period with an emphasis on the relationship between training load and dietary intake. Moreover, we also propose a Carbohydrate Periodization Index (CPI), a single metric to capture how tightly an athlete’s carbohydrate intake is adjusted based on their training load, the magnitude of adjustment, and how frequently these adjustments occur.

## 2. Methods

### 2.1 Study design

This observational study monitored the daily self-reported and self-selected nutrition intake and exercise training of endurance athletes for 12 weeks. Throughout the study period, participants were free to perform any type of training or racing and consume any type of diet, provided it was tracked appropriately. In addition to diet and exercise, measures of sleep, heart rate variability (HRV), and subjective wellbeing were recorded daily. Data presented herein are from a wider study of endurance training and recovery and are focused on the relationships between dietary intake and training load. The study was open to male and females aged 18 or older who train at least seven hours per week, use a smartphone app to track their dietary intake at least five days per week, capture HRV daily, and track sleep using a wearable device. All study protocols and materials were approved by the Auckland University of Technology Ethics Committee (22/7).

### 2.2 Participants

The study was completed by 55 endurance athletes (61.8% male, aged 42.6 ± 9.1 years, training 11.6 ± 3.9 hours per week) from 10 countries (Supplemental Fig. 1). The primary sports represented were triathlon (67.3%), running (20.0%), cycling (10.9%), and rowing (1.8%). The self-reported competitive level included professional (2.6%), elite non-professional (qualify and compete at the international level as an age-group athlete, 34.6%), high-level amateur (qualify and compete at National Championship-level events as an age-group athlete, 25.6%), and amateur (enter races but don’t expect to win, or train but do not compete, 37.2%) athletes.

**Figure 1.**
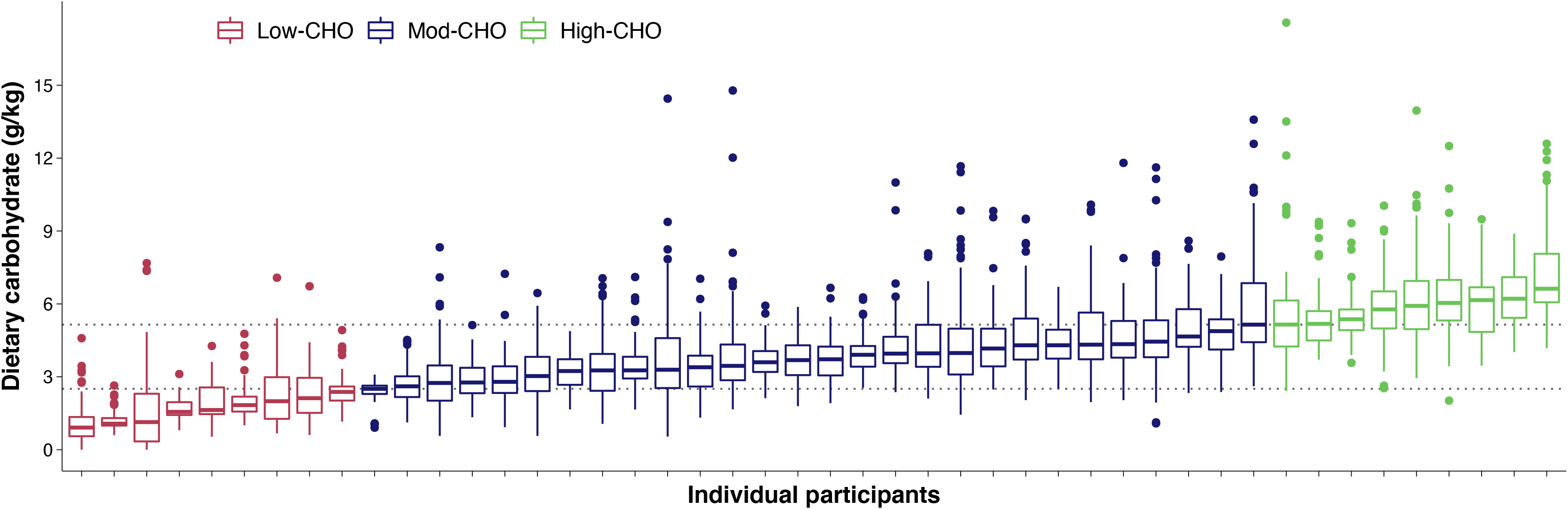
Box plot of daily carbohydrate (CHO) intake for each participant, color coded by habitual dietary intake defined as a median daily CHO intake < 20th percentile (2.5 g/kg, Low-CHO), > 80th percentile (5.1 g/kg, High-CHO), and between these values as moderate (Mod)-CHO

### 2.3 Assessment of self-reported training load

All exercise was recorded in Training Peaks software (TrainingPeaks, Louisville, CO, USA). Each session was noted for modality (e.g., bike, run, swim), total time, and session rating of perceived exertion (sRPE [16]) using the Borg CR100® scale, which offers additional precision compared with the CR10 scale [17–19]. Participants were asked to rate the session within 1-h of exercise, but sRPE scores are temporally robust from within minutes to within days following a bout of exercise [16]. Additionally, participants noted the amount of carbohydrate consumed within the 4-h pre-exercise window.

### 2.4 Assessment of self-reported dietary intake

Participants were instructed to maintain their typical dietary habits and record all calorie-containing food and drink consumed for the duration of the 12-week study. Weighing of food was encouraged, but not mandated, and common issues such as underreporting were discussed prior to beginning the study. Participants were not required to record fluid ingestion, micronutrient content of foods, or the timing of meals (apart from the pre-exercise carbohydrate amount). Dietary intake was self-reported via the MyFitnessPal application (www.myfitnesspal.com) [20, 21]. Compliance with dietary tracking was monitored by connecting to participant food logs via MyFitnessPal, and enquiring about any unexpected values (determined both visually and using anomaly detection software [22]). Incomplete days of tracking (2.2 ± 4.6% of days per participant) were removed from the data. To aid compliance, participants were recruited who were already regularly tracking their diet (in several cases daily for 4+ years), and so all participants displayed strong intrinsic motivation for habitual diet tracking. As an additional check of compliance, analysis of the calorie intake trend over time was performed for each participant (described in section 2.5).

### 2.5 Data analysis

Training load was calculated for each workout as the product of sRPE and duration of exercise in minutes [23], divided by 10 to account for the 100-point scale. Exercise was summed into daily totals for workout duration and training load, along with coded variables for modality of workout (e.g., swim, bike, run, strength, other) and if any training was performed in the fasted state. Because dietary protein and fat ingestion have minimal effects on substrate oxidation [24, 25], fasted training was defined as consuming < 5 g of carbohydrate in the 4-h pre-exercise window. For multiple exercise sessions in a single day, a weighted mean based on the duration of each session was used to calculate a single value for pre-exercise carbohydrate ingestion in grams. External load metrics such as heart rate (HR), power, or pace were not collected because many athletes undertake activities that can’t be quantified on a common scale such as strength training, yoga, or swimming without a HR monitor, and also because the sRPE is considered to be a valid and reliable method for calculating training load across modalities [23].

Dietary macronutrient intake was calculated relative to body mass (g/kg) to compare across participants more appropriately. Because the beliefs and practices relating to pre-exercise nutrition vary greatly based on habitual diet patterns [12], participants were classified into three sub-groups based on percentile of median daily carbohydrate intake. Low-carbohydrate was considered < 20th percentile (2.5 g/kg, Low-CHO), high-carbohydrate was considered > 80th percentile (5.1 g/kg, High-CHO), and moderate-carbohydrate was between low and high (Mod-CHO). These values fall in line with commonly used delineations [1], and allow comparisons to be made between sub-groups.

As an additional check of dietary compliance, analysis was performed to determine any negative trend in reported daily calorie intake that could not be explained by changes in training load or body weight. If someone’s reported intake gradually decreases over time it could indicate waning compliance or attention to detail in dietary tracking. A time series that does not exhibit any trend and displays constant variance and constant autocorrelation over time is referred to as stationary [26]. For each participant, stationarity of the time series for daily calorie intake was checked using the Kwiatkowski-Phillips-Schmidt-Shin test [27]. If non-stationarity was found, the Breusch-Pagan test [28] was performed to determine if the non-stationarity could be due to changing variance over time (heteroscedasticity), and the Ljung-Box test [29] was performed to determine the presence of autocorrelation. If a participant’s calorie intake was found to be non-stationary but did not display heteroscedasticity or autocorrelation it was assumed to have a significant trend. To account for potential changes in training load and/or body weight, linear models were created for each participant presenting a trend, modeling daily calorie intake as a function of study day, exercise load, and body weight, with 95% confidence intervals for model coefficients generated from 1000 bootstrap resamples [30]. If the confidence intervals did not cross zero, the participant was assumed to have a significant trend in reporting that was not due to changes in exercise load or body weight and was excluded from analysis if the coefficient for study day (after adjusting for exercise load and body weight) was ≤ −5. This value was chosen to allow for unaccounted variations in non-exercise activity in response to changes in meal patterns, which can influence daily energy requirements [31]. At least 85% of a participant’s training and diet must have been logged for data to be included in the analysis. Missing values were imputed using multiple linear regression [30, 32]. Participants training on average less than 6 h per week were also excluded from analysis, due to the lack of significant exercise stress. Participants who did not complete the full 12 weeks due to illness, injury, or drop-out but completed at least 6 weeks of tracking were included in the analysis (n = 12).

### 2.6 Carbohydrate Periodization Index

A carbohydrate periodization index (CPI) was created as a single value that could capture the combination of how closely an athlete modulates their carbohydrate intake based on training load, the degree of day-to-day variation in carbohydrate intake, and how frequently these modulations occur, as:

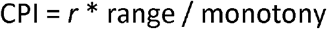

Where *r* represents the Pearson correlation value between daily training load and daily carbohydrate intake (g/kg). Range is calculated as the difference between the highest and lowest single-day carbohydrate intake (g/kg). To quantify day-to-day variability in carbohydrate intake, monotony is calculated as average daily carbohydrate intake (g/kg) divided by the standard deviation, based on the concept of the training monotony score [33]. Simulated CPI values are shown in Supplemental Fig. 2, across a range of inputs. Based on the data from this study, as well as published [13] and unpublished data from elite athletes, we interpret CPI scores using the following guidelines:

< 1.0: no evidence of dietary carbohydrate periodization
1.0 – 2.0: evidence of moderate dietary carbohydrate periodization
2.0 – 3.5: evidence of a high degree of dietary carbohydrate periodization
> 3.5: evidence of extreme dietary carbohydrate periodization

**Figure 2.**
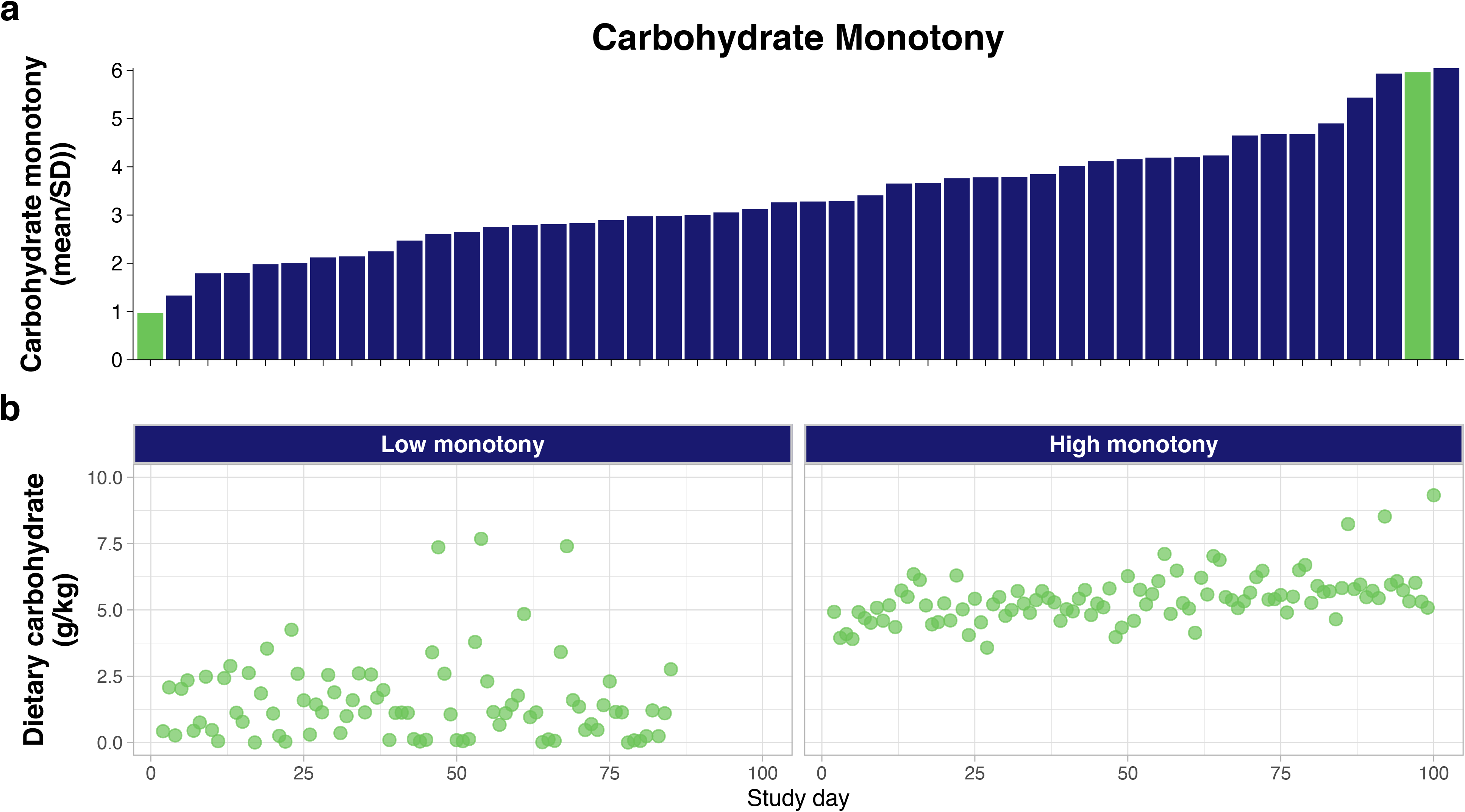
Carbohydrate monotony scores for each participant (a), and example data from participants with low and high monotony scores (b). Green bars in (a) correspond to the data shown in (b). For (b), the x-axis represents the sequential days of the study tracking period

### 2.7 Sub-group analysis

For sub-group analysis, professional athletes (n = 2) were merged into the elite non-professional group. A series of linear models were used to estimate differences in CPI, weekly training volume (hours per week), and percentage of training days that included fasted-state training based on habitual carbohydrate intake, sex, and competitive level. Contrasts within each sub-group (e.g., diet, competitive level, or sex) were estimated after adjusting for the other variables using the *emmeans* R package, with multiple comparisons adjusted using the Holm correction. A standardized effect size for each contrast was computed and interpreted as small (0.2), medium (0.5) and large (0.8) [34].

To visualize the relationship between carbohydrate intake and training load, exercise duration, and exercise intensity, univariable regression analysis was performed separately for each habitual diet group, with the best-fit regression line (linear or polynomial) established using the likelihood ratio test. Because multiple data points were collected from each participant, the assumption of independence of residuals is violated. Therefore, we built general linear mixed-effect models to examine how factors were related, specifying subject ID as a random intercept using the *lme4* R package [35], and report the marginal R, which describes the proportion of variance explained by only the fixed effect. All models were checked by visualizing the Q–Q and other residual plots to ensure approximate residual normality and heteroscedasticity, using the performance R package. Descriptive statistics are provided as mean ± SD. All analyses were carried out with R version 4.0.3 (The R foundation for Statistical Computing, Vienna, Austria), with the level of significance set at p < 0.05.

## 3. Results

### 3.1 Daily Carbohydrate intake

After removing participants training less than 6 h per week (n = 5) and those displaying a downward trend in dietary reporting that could not be explained by training load or body weight (n = 4), the final analysis included 46 participants (60.9% male) and 3,718 days of dietary tracking (80.8 ± 11.7 days per participant). Boxplots of carbohydrate intake for each participant are shown in Fig. 1, colored by habitual dietary intake. Mean daily carbohydrate intake was 3.9 ± 1.5 (range 1.2 to 7.2) g/kg, with the highest single-day intake being 17.6 g/kg. Exercise was performed in the overnight-fasted state on 27.2 ± 22.0 (range 0 to 80.6) % of training days. Daily carbohydrate intake for each participant is shown in Supplemental Fig. 3, along with an indication of sessions performed in the overnight-fasted state.

**Figure 3.**
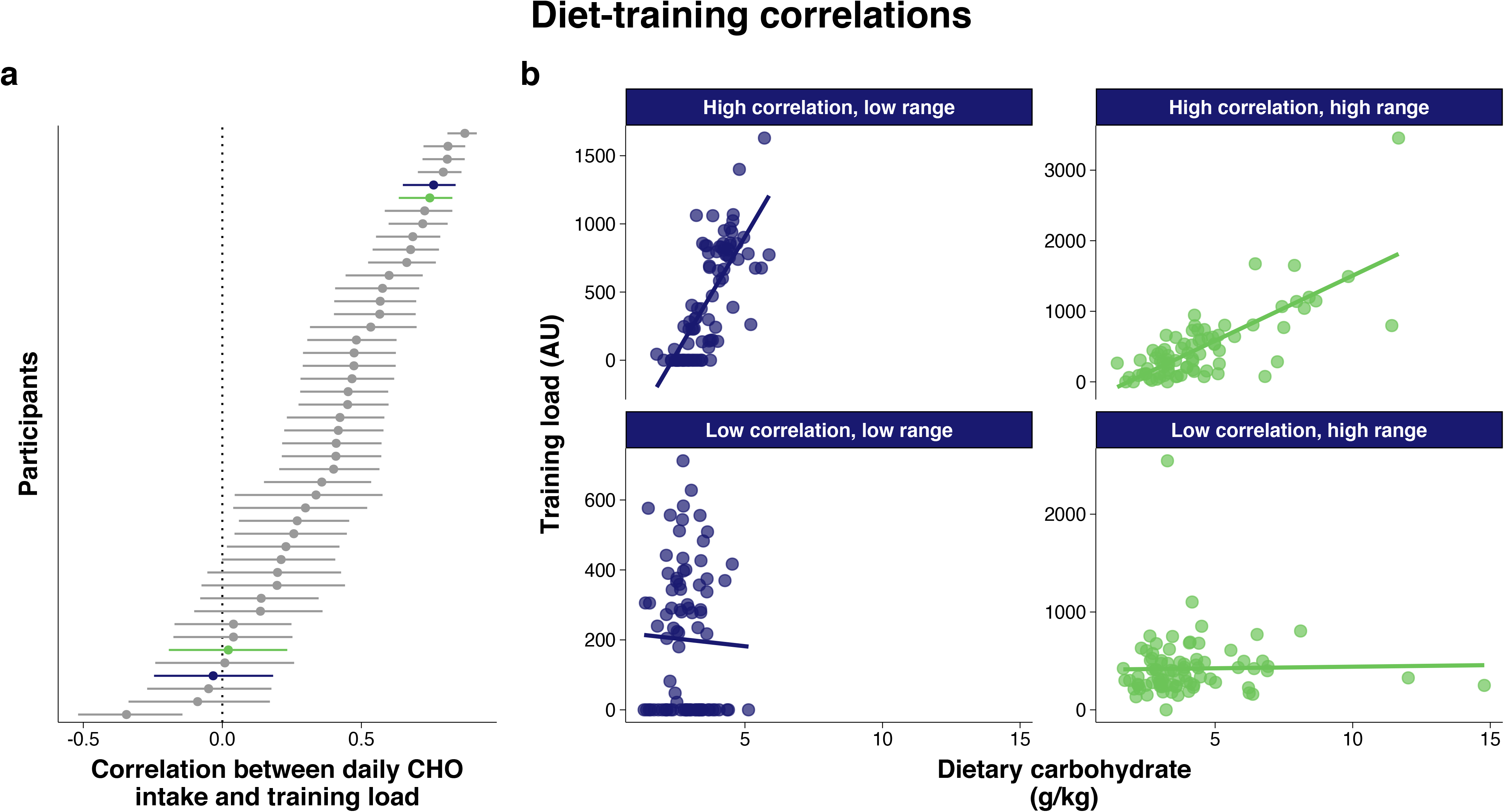
Pearson correlation values with 95% confidence intervals are shown between training load and daily carbohydrate (CHO) intake (g/kg) for each participant (a), and example data from participants with combinations of low and high correlations and carbohydrate ranges (b). Colored bars in (a) correspond to the data shown in (b). Training load is the product of session rating of perceived exertion and duration in minutes. Range refers to the difference between an athlete’s highest and lowest daily carbohydrate intake and can be inferred in (b) as the horizontal distance of the trend line. AU: arbitrary units

### 3.2 Carbohydrate Periodization Index

The CPI encompasses the range of daily carbohydrate intake, carbohydrate monotony, and the correlation between carbohydrate intake and training load. The mean range of daily carbohydrate intake (difference between the highest and lowest days for each participant) was 6.6 ± 3.1 (range 2.0 to 15.2) g/kg. Carbohydrate monotony values ranged from 1.0 to 6.0 and are shown in Fig. 2, along with example participant data of low- and high-monotony diets. Pearson correlations between training load and daily carbohydrate intake (g/kg) ranged from - 0.34 to 0.87, and are shown in Fig. 3, along with examples of participants with high and low correlation values and high and low carbohydrate ranges. Correlation plots for all participants are shown in Supplemental Fig. 4. The CPI scores for each participant are shown in Fig. 4, along with examples of participants with high and low scores. The median CPI score was 0.6 (range - 1.2 to 5.6), with 34.8% of participants attaining a CPI of at least 1.0 and just 15.2% of participants attaining a CPI of at least 2.0.

**Figure 4.**
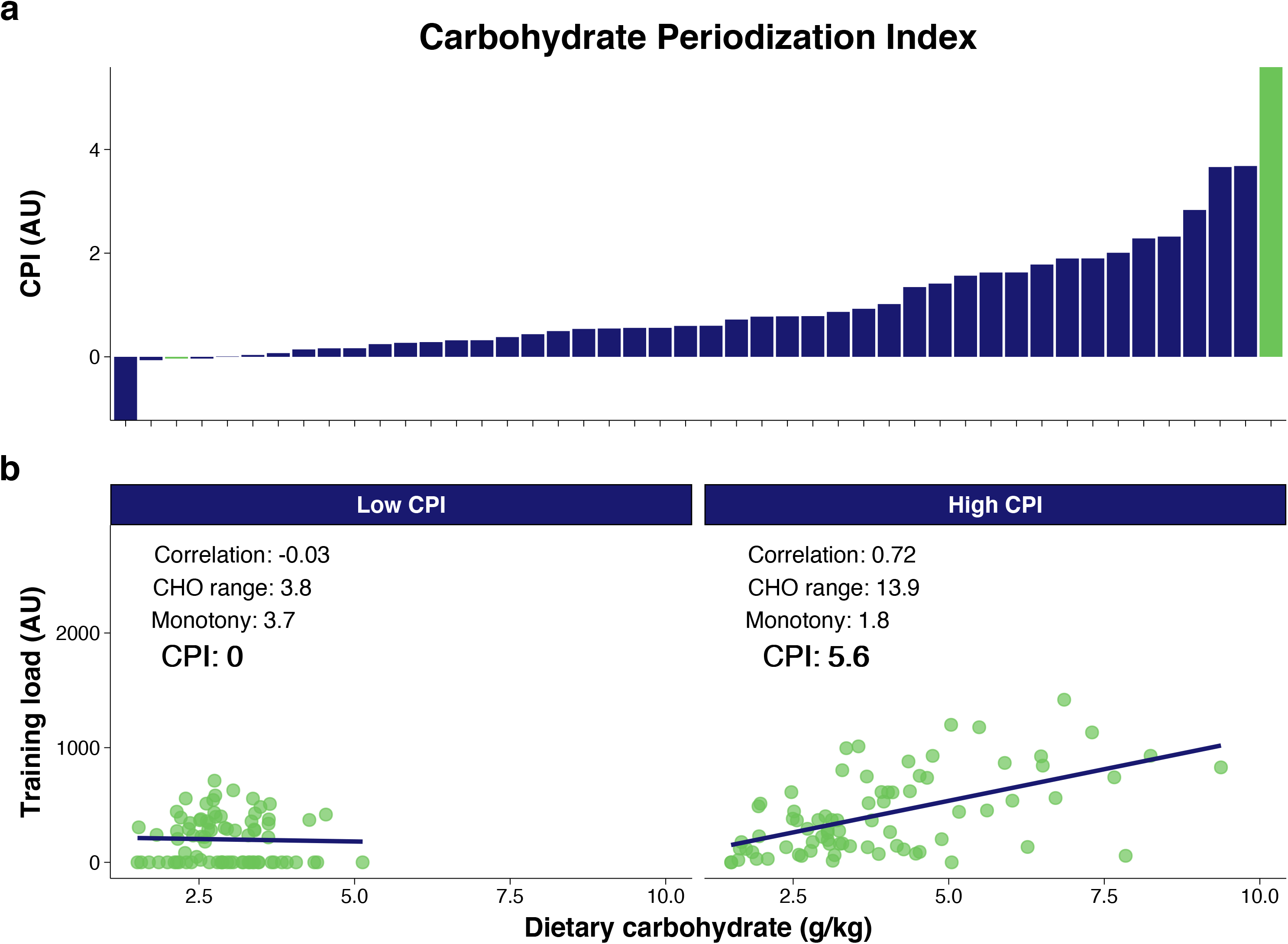
Carbohydrate Periodization Index (CPI) scores for each participant (a), and example data from participants with low and high scores (b). Colored bars in (a) correspond to the data shown in (b). Training load is the product of session rating of perceived exertion and duration in minutes. AU: arbitrary units, CHO: Carbohydrate

### 3.3 Sub-group analysis

The CPI values were higher (p < .05) among the highest-level compared with amateur athletes, but not different based on habitual diet or sex (Fig. 5). The percentage of training days an athlete performed fasted training was higher (p < .05) for Low-CHO (42.5 ± 22.5% of training days) compared with High-CHO (13.4 ± 14.1% of training days), and for males (33.6 ± 24.0% of training days) compared with females (17.2 ± 13.9% of training days), but there were no differences based on competitive level (Supplemental Fig. 5). Weekly training volume was not different based on habitual diet, competitive level, or sex (Supplemental Fig. 6).

**Figure 5.**
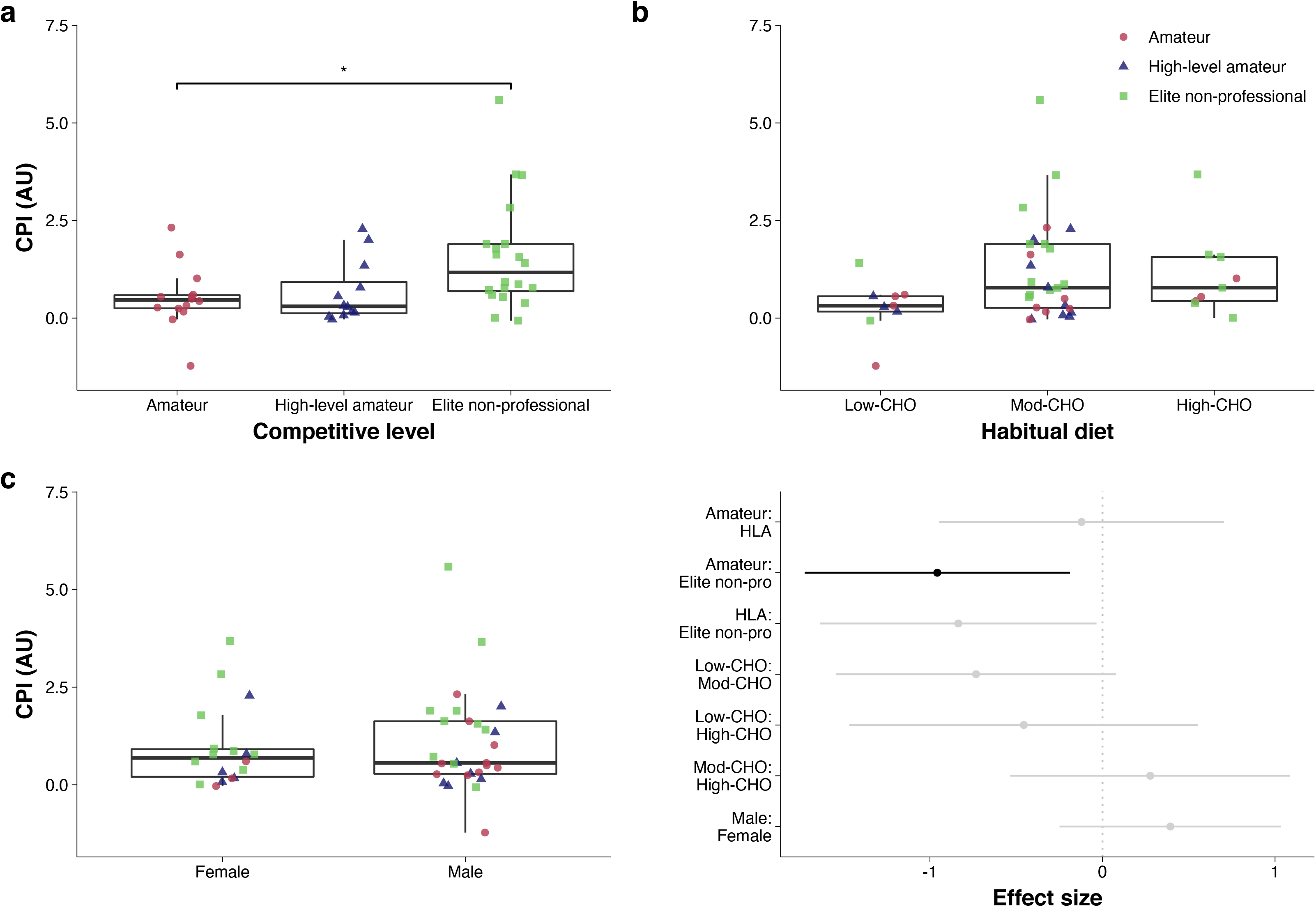
Boxplot of Carbohydrate Periodization Index (CPI) values, separated by competitive level (a), habitual dietary pattern (b), and sex (c), and effect sizes with 95% confidence intervals for all pairwise contrasts (d). Individual participants are shown separated by shape and color based on competitive level. * indicates p < 0.05, effect sizes shown in black correspond to pairwise comparisons with significant p-values after adjusting for multiple comparisons. AU: arbitrary units

### 3.4 Diet-exercise adjustment

Daily carbohydrate intake in relation to training load (product of duration and sRPE), exercise duration, and sRPE is shown in Fig. 6a–c. The stronger relationships seen between exercise duration and daily carbohydrate intake, compared with exercise RPE and daily carbohydrate intake, indicate athletes are more responsive with their diet to changes in exercise duration than intensity. However, Low-CHO athletes displayed very little relationship between dietary carbohydrate and training. Carbohydrate intake during the 4-h pre-exercise window is shown in Fig. 6d–f in relation to the subsequent training load, duration, and intensity, with evidence of weak relationships between pre-exercise carbohydrate and the subsequent exercise duration and intensity (Fig. 6d–f). Daily carbohydrate intake in relation to the day before, day of, and day after a given training load is shown, separated by habitual diet, in Supplemental Fig. 7. A relationship is seen between daily carbohydrate intake and training load on the day of exercise for Mod-CHO and High-CHO groups, but not the day before or after a given training load for any sub-groups.

**Figure 6.**
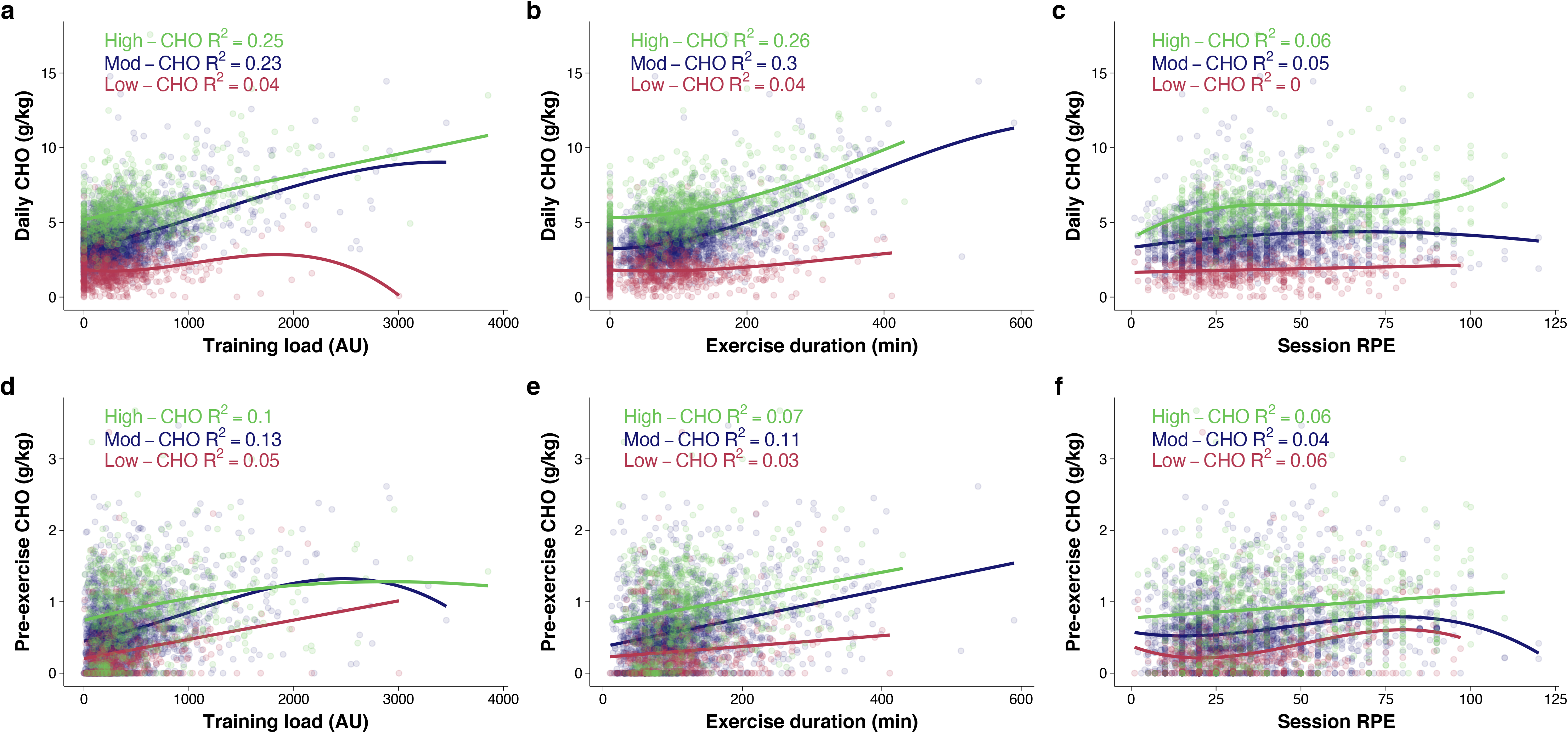
Daily carbohydrate (CHO) intake relative to training load (a), exercise duration (b) and exercise intensity as rating of perceived (RPE, c), and carbohydrate intake within the 4-h pre-exercise window in relation to training load (d), exercise duration (e), and RPE (f), separated by habitual diet. Training load is calculated as the product of session RPE and exercise duration in minutes, divided by 10. Best-fit regression lines based on univariable linear mixed effects models are shown for each diet group, with fit indicated as R. AU: arbitrary units

## 4. Discussion

This study provides novel data on the self-selected dietary intake of endurance athletes acros a 12-week period, highlighting the relationships between training load and dietary carbohydrate. The primary findings are that many endurance athletes are not following current sport nutrition guidelines to periodize carbohydrate intake based on training load, or if they are, the magnitude of adjustment is small. We also introduce a Carbohydrate Periodization Index (CPI), a single metric to capture the combination of how tightly an athlete’s carbohydrate intake is adjusted based on training load, the magnitude of adjustments, and how frequently these adjustments occur. The CPI can serve as a tool for researchers, coaches, and athletes to quantify carbohydrate periodization practices, discern changes in intake patterns across training cycles, and make comparisons within and between individuals.

Prior to this study, knowledge of the day-to-day dietary periodization practices of athletes has largely been limited to questionnaires [9–12], case studies [13, 14], and short-duration (∼7-d) observations of endurance [15] and team-sport [36–38] athletes. Longer-duration observational studies can help answer questions that would not be feasible to study in a controlled laboratory environment. To this end, we recruited athletes who were already regularly, and voluntarily, tracking their diet and training so we could obtain longer-term longitudinal data across a variety of training regimens. Although dietary intake in the general population is likely to be under-reported [39], this would not necessarily apply to a population of athletes who are highly motivated and already in the habit of tracking dietary intake. It has been suggested that familiarity with and interest in keeping food records may lead to more reliable estimates of energy intake [40]. Furthermore, in the context of this study within-participant comparisons are more important than between-participant comparisons, minimizing the influence of variation in dietary tracking among participants.

### 4.1 Dietary Carbohydrate Intake

Participants displayed a wide range of daily carbohydrate intakes (Fig. 1). We observed an overall pattern of carbohydrate intake that increased with training loads, regardless of habitual diet, that appears to be primarily attributed to athletes adjusting their intake in response to exercise duration rather than exercise intensity (Fig. 6a–c). Positive, yet weaker, relationships can also be seen between pre-exercise carbohydrate intake and subsequent training load and duration (Fig. 6d–f). From this perspective, athletes are generally following the “fuel for the work required” framework [3]. However, at the individual level the relationship between daily training load and daily carbohydrate intake varied greatly (Fig. 3). Similarly, at the group level athletes trained in the overnight-fasted state roughly one-quarter of training days, but at the individual level this ranged from 0–81 % of training days. The lack of generalizability from group to individual responses is known as nonergodicity [41], and highlights the need for individualized analysis of carbohydrate periodization practices.

### 4.2 Carbohydrate Periodization Index

The CPI provides a single number to capture multiple facets of the diet-exercise interaction and offers a unifying framework to compare periodization practices among athletes. There are three components of the CPI — correlation, range, and monotony. The correlation between carbohydrate intake and training load offers insight into how tightly an athlete links their intake with their training, and the range of daily carbohydrate intake provide a measure of how extreme the changes are between high- and low-volume training days. It is also of interest to know how frequently these changes in diet and training occur. For example, someone could very closely match their intake and training, yet perform very similar types of training that wouldn’t warrant large changes in carbohydrate intake. Based on the concept of the training monotony score [33], we calculated a carbohydrate monotony score to quantify day-to-day variability in carbohydrate intake (Fig. 2). This number reflects the relationship between average daily carbohydrate intake and the standard deviation, meaning it decreases as the daily variation in carbohydrate intake increases.

Being a composite of three variables, one could achieve the same CPI score in a variety of different ways (Supplemental Fig. 2). Therefore, at the individual level interpretation should be informed by the context of the athlete’s training load (e.g., large training volumes would be needed to achieve a high range of carbohydrate intake), habitual dietary pattern (e.g., lower-carbohydrate athletes may struggle to achieve a high range), and personal preferences. The length of observation period could also influence the achievable CPI values. A very high CPI (> 3.5) is more likely attained during a short period of time such as a cycling Grand Tour, where the energy expenditure can vary greatly across stages and rest days. This allows for a large range of carbohydrate intake within a short time, lowering the monotony score. Additionally, few recreational athletes could achieve the workload of professional cyclists, particularly during a Grand Tour, thereby placing a *de facto* limit on their available carbohydrate intake range. While there is no theoretical ceiling, a combination of the most extreme values observed in our data for carbohydrate range (15.2 g/kg) and correlation (0.87) and monotony (1.0), would yield a CPI of 13.2. However, these values seem unlikely to be attained in any practical scenario.

The CPI requires a measure of training load to determine the correlation between carbohydrate intake and training load. Training load can be measured and classified as either internal and/or external, based on the measurable aspects occurring internally or externally to the athlete [42]. Internal load reflects the relative physiological strain and disturbance in homeostasis of the metabolic processes in response to an external load, which is characterized by objective measures such as distance, power, or speed [43, 44]. Internal load has been recommended as the primary measure when monitoring athletes, as it plays a pivotal role in determining training outcomes and can reflect variations in the stress response to a given external load due to other stressors such as extreme temperature, or accumulated training fatigue [42]. The sRPE-based load metric we used provides a measure of internal load, and was chosen as a valid and reliable method for calculating training load across exercise modalities [23], a necessity with multi-sport athletes. Our study used the CR100® scale, which uses ratio scaling to account for the non-linear stimulus-response relationship and overcomes many limitations of other scales [17–19, 45]. For example, the CR100® places ‘Maximal’ at 100, but the scale goes to 120. This is because ‘Maximal’ is anchored in a previously experienced strongest intensity (i.e., the hardest someone has ever gone), but for some things such as sensations of pain, an absolute maximum can be at a level above this. To avoid a ceiling effect, the “absolute maximum” is placed with a dot outside the given numerical scale [17]. Because the CPI is based on the correlation between dietary intake and training load rather than any absolute load values, the use of external load measures such as exercise energy expenditure or watts for cyclists, or GPS-based metrics in team sports could also theoretically be used. Indeed, analysis of a cyclist in this study who performed nearly all training on a bike with a power meter revealed very similar CPI scores when basing it on training load (0.7), total work done during training (0.6), or the Training Peaks composite Training Stress Score [46] (0.8). This is in accordance with the very large correlations reported across multiple measures of training load in cyclists during racing and training [47]. However, future research is needed to perform a more rigorous examination of the variation in CPI when calculated using different measures of training load before they can be used interchangeably.

Using data from this study along with unpublished observations, we propose cutoff values for interpretating the CPI (see methods) and found only about one-third of athletes in this study displayed even moderate levels of periodization based on these values. This is a finding that should be generalizable to the wider population of endurance athletes as study participants were not pre-screened based on any dietary practices nor were they told that the diet-training relationship would be analyzed in this manner. However, by habitually tracking their diet and training it is possible these participants could adjust their diet based on training volume more easily and precisely than someone not tracking their diet.

Another reason for creating the CPI is that many athletes who are aware of the sport nutrition recommendations for modulating carbohydrate with exercise may be unclear if or how they are implementing them. A study in elite runners and race-walkers compared dietary intake between hard and easy days of training, and found only modest evidence of periodized nutrition practices among females (6.2 vs. 5.8 g/kg carbohydrate on hard vs. easy days, respectively), and no differences among males (7.3 vs. 7.2 g/kg carbohydrate on hard vs. easy days, respectively), despite the majority of athletes reporting an understanding of dietary periodization practices [15]. Furthermore, there were no differences in pre-exercise carbohydrate intake before hard or easy workouts, or between sexes [15]. Our data also reveal a similar degree of carbohydrate periodization among males and females, but a wider range of carbohydrate ingestion across days, likely owing to the longer duration and greater heterogeneity of subjects in our study.

### 4.3 Sub-group analysis

We previously reported that beliefs and practices relating to pre-exercise nutrition vary greatly based on sex, competitive level, and habitual dietary pattern [11, 12]. Because of this, we analyzed the data by sub-groups and report higher CPI scores among the highest-level athletes. This is in accordance with survey results showing the percentage of athletes who reported following a periodized-carbohydrate diet increased with competitive level from 8% of amateur athletes to 17% of elite non-professional and 27% of professional athletes [12]. It was also found that Low-CHO performed a greater percentage of their training sessions in the overnight-fasted state (Supplemental Fig. 5), which is also in accordance with data collected via survey [11]. It should be noted that the dietary comparisons are inherently challenged by the loss of fidelity that comes from combining a continuous variable into three groups. For example, athletes with very similar carbohydrate intakes on either side of the cutoff values get placed into different groups. It was unexpected to find no differences in weekly training volume based on competitive level (Supplemental Fig. 6). However, this may be due to the time of year the study took place (off-season in the Southern hemisphere) and some amateur athletes with unusually high training volumes.

### 4.4 Limitations

There are several limitations to this study, and to the CPI. Most notably, the use of athlete self-report dietary data. Although athlete populations have been shown to underreport dietary intake [48], nearly all previous studies have used short-duration food records rather than smartphone apps. Smartphone-based food diaries lead to better participant compliance, compared with paper-based diaries [49], possibly related to the additional features such as the ability to scan barcodes on food packages and save commonly-consumed foods. It has also been suggested that familiarity with and interest in keeping food records may lead to more reliable estimates of energy intake [40]. High reliability has been reported with food-tracking apps [50], and in the context of this study it could be argued that reliability is of greater importance than validity. Moreover, the ecological validity of our study is high, as this is the same type of data that would be received by coaches and nutritionists working with athletes.

A limitation related to the CPI is its inability to discern within-day carbohydrate manipulation, such as twice-daily training without carbohydrate between sessions [6] or an overnight sleep-low approach [51, 52]. However, aligning with the “fuel for the work required” paradigm [3], appropriate daily intake should still be ensured regardless of the within-day timing. Future work could consider a within-day index to work alongside or as part of the CPI. Athletes following a lower-carbohydrate diet would typically have a lower range of carbohydrate intake, which could lead to lower CPI values despite a conscious effort to manipulate intake. An additional limitation is that the correlation values could be influenced by outliers, in which case an alternative to the Pearson calculation such as the Spearman correlation might be more appropriate [53]. Although we considered adjusting the correlation coefficient for each participant based on the distribution and existence of outliers, we decided against this because outlier values matching a very large training load are both appropriate and important, in the context of fueling for the work required. Furthermore, beyond its utility in the research setting the CPI was created as a practical tool for athletes and coaches. Therefore, the required calculations should be easily performed on widely available spreadsheet software, without needing to calculate and choose from multiple correlation coefficients.

### 4.5 Future Directions

Future research can determine if our results are generalizable to a wider population of endurance athletes, if CPI values are robust to different training load metrics, if normative values are comparable across various endurance sports and competitive levels, and how the CPI can be applied outside of endurance sports. It would also be valuable for future carbohydrate periodization studies to report the CPI to quantify differences in intake patterns between groups (e.g., studies comparing chronically high- or low-carbohydrate with periodized carbohydrate intake). This could allow the CPI to be used as a moderator in meta-analyses, which have thus far found no effect on performance from periodized carbohydrate intakes but have lacked a way to quantify the periodization [54].

## 5. Conclusion

This study utilized a novel approach to monitoring endurance athletes throughout 12 weeks of self-selected training to better understand the current real-world application of dietary periodization. We show that many endurance athletes are not following current sport nutrition guidelines to periodize carbohydrate intake based on training load, or if they are, the magnitude of adjustment is small. We have also introduced the CPI, a single metric that can be used by researchers, coaches, and athletes to quantify carbohydrate periodization practices, as well as make comparisons within and between individuals and, potentially, across populations.

## Data Availability

The R code used in this analysis is publicly available at (https://github.com/Jeffrothschild/CPI_code).

## Author contributions

JAR conceptualized and performed the analyses and wrote the first draft of the manuscript. All authors edited and revised the manuscript and approved the final version of the manuscript.

## Acknowledgements

A preprint of this manuscript has been posted on medRxiv

