## supplemental figs for "The quantification of daily carbohydrate periodization among endurance athletes during 12 weeks of self-selected training: presentation of a novel Carbohydrate Periodization Index"

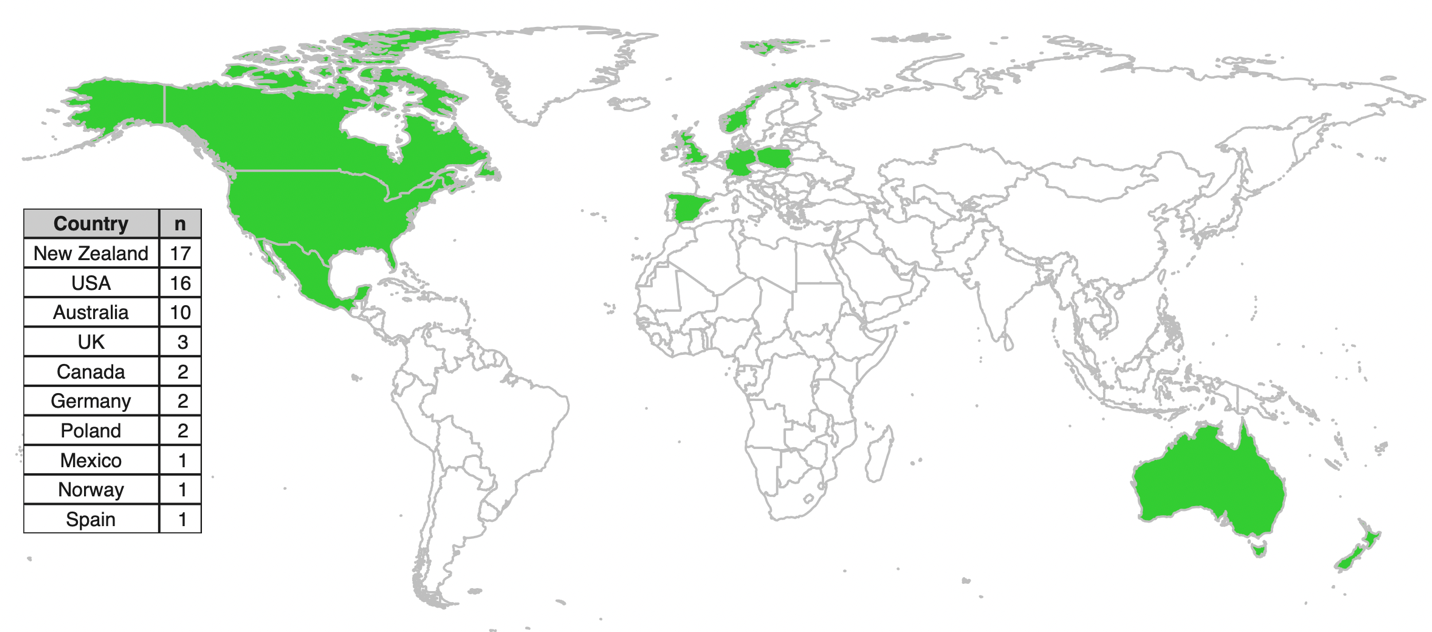

**Supplemental Fig. 1** Location of study participants

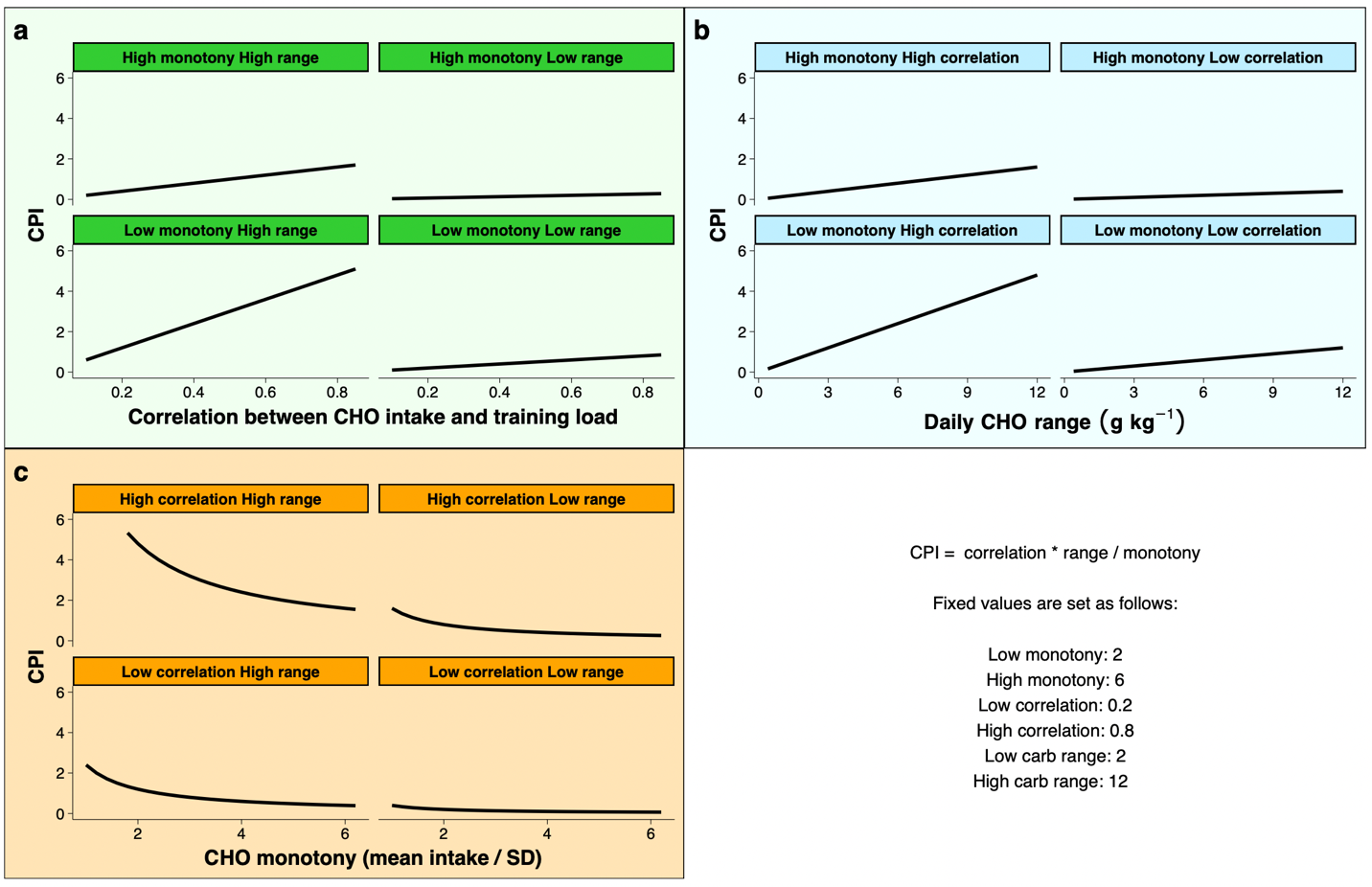

**Supplemental Fig. 2** Simulated CPI values across a range of correlations (a), carbohydrate (CHO) intake ranges (b), and CHO monotony values (c).

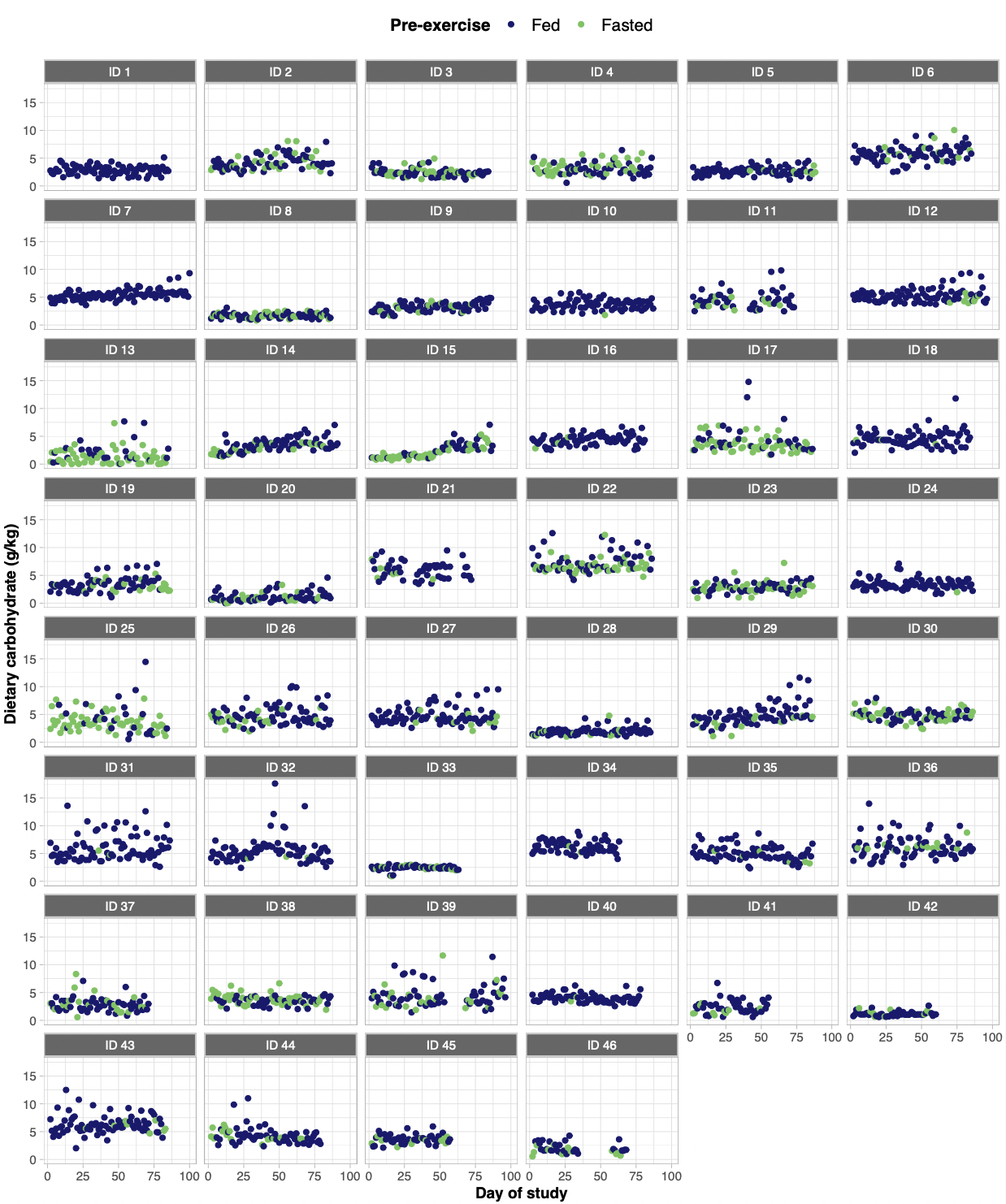

**Supplemental Fig. 3** Daily carbohydrate intake (g/kg) for each day of the study, separated by participant and color-coded according to if training was performed in the overnight-fasted (light green circles) or fed (dark blue circles) state

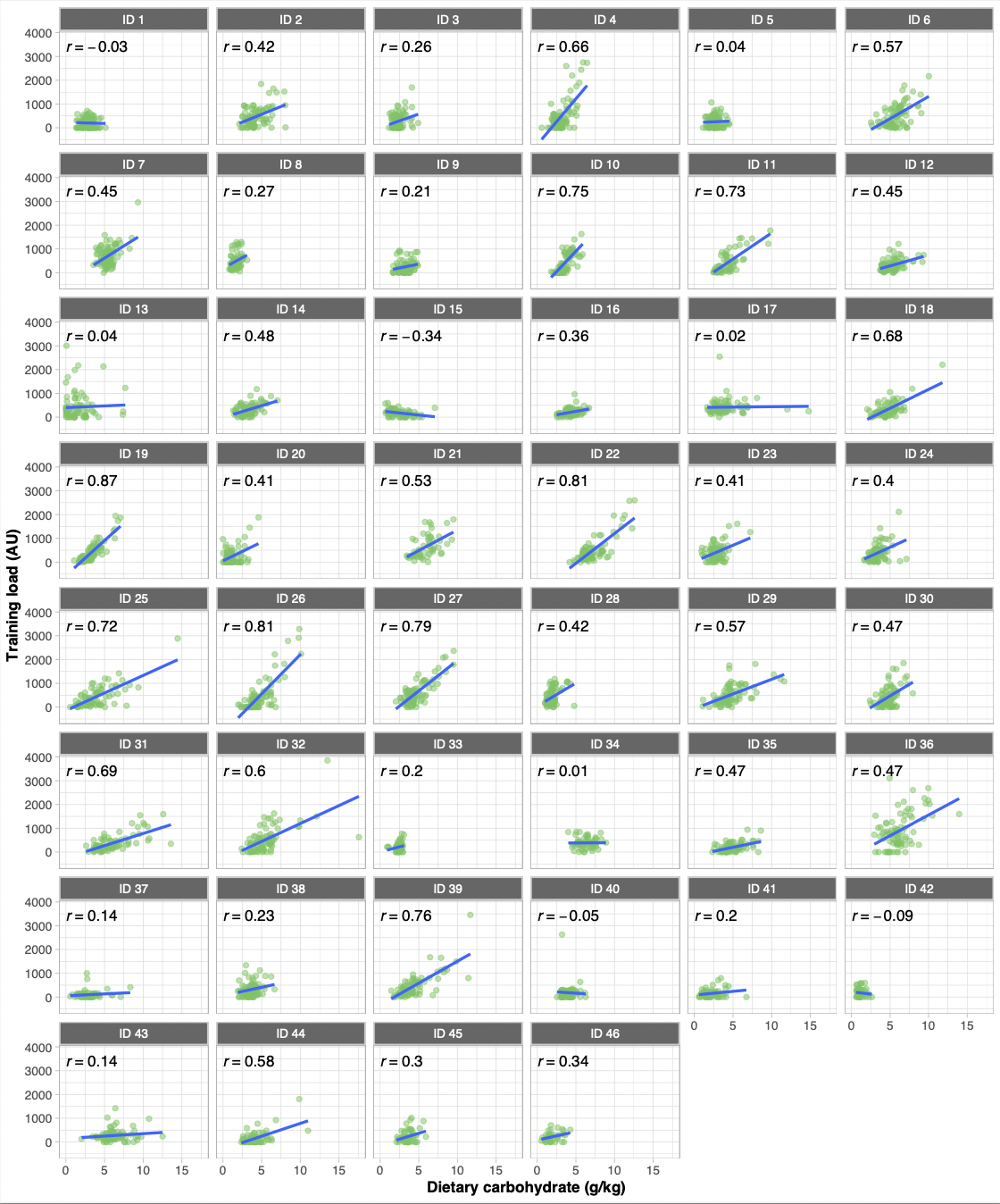

**Supplemental Fig. 4** Relationships between daily carbohydrate intake (g/kg) and training load (product of session RPE and duration), for each participant. Pearson correlations are shown for each participant.

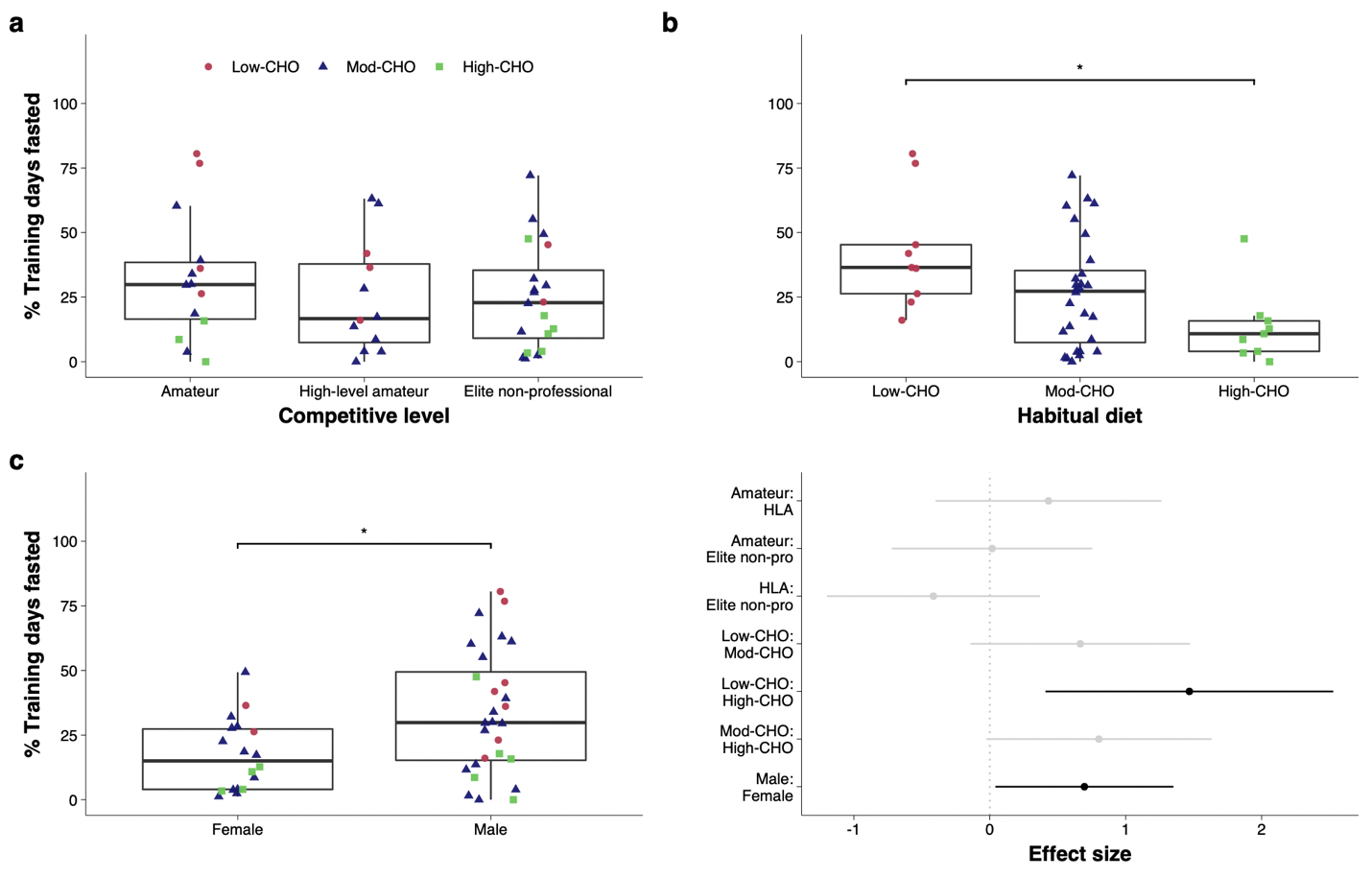

**Supplemental Fig. 5** Boxplot of percentage of days training in the overnight-fasted state, separated by competitive level (a), habitual dietary pattern (b), and sex (c), and effect sizes with 95% confidence intervals for all pairwise contrasts (d). Individual participants are shown separated by shape and color based on dietary sub-group. * indicates p < 0.05, ** indicates p < 0.01, effect sizes shown in black correspond to pairwise comparisons with significant p-values after adjusting for multiple comparisons

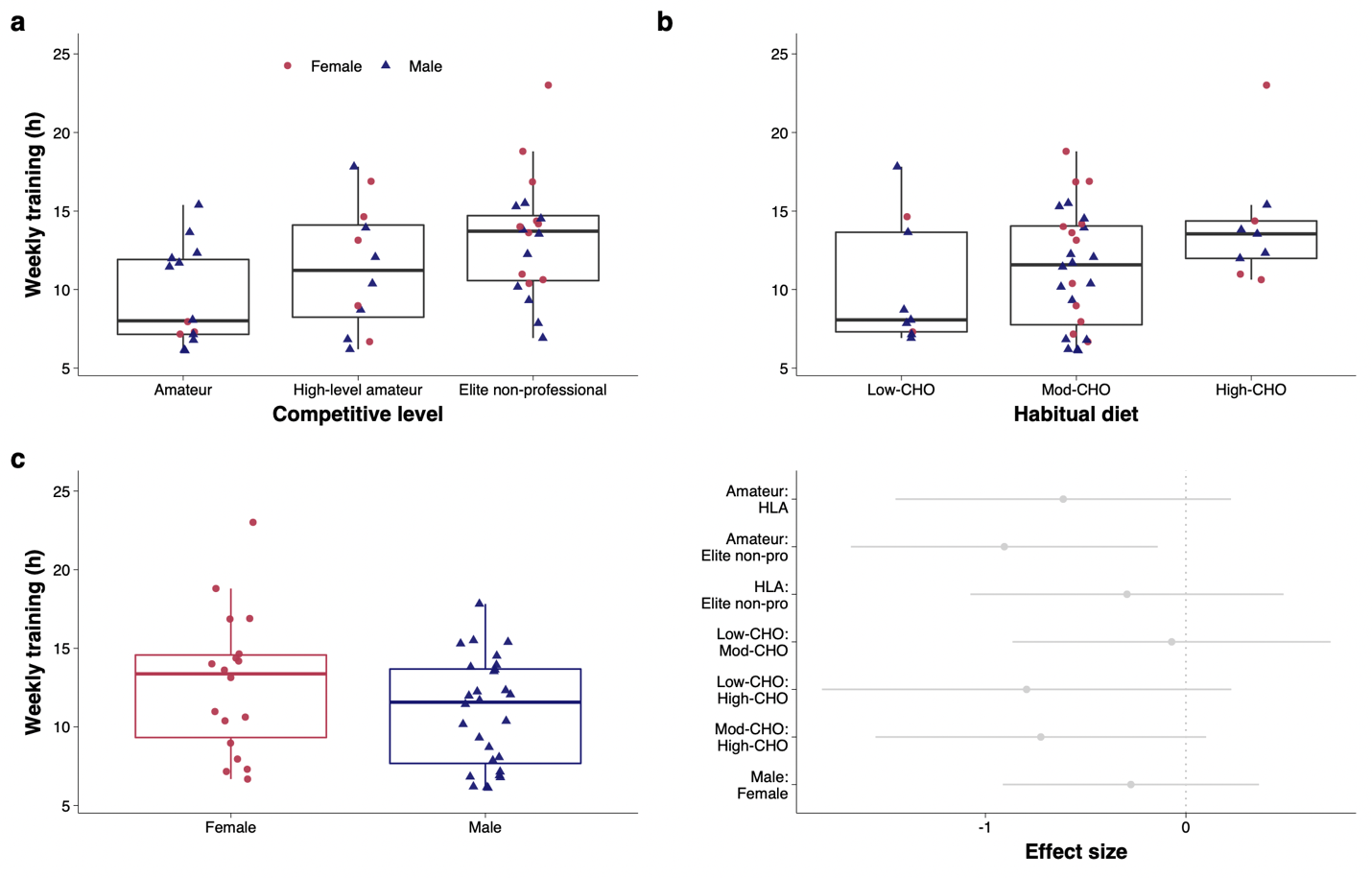

**Supplemental Fig. 6** Boxplot of training volume, separated by competitive level (a), habitual dietary pattern (b), and sex (c), with Tukey’s post-hoc comparison tests. Individual participants are shown separated by shape and color. * indicates p < 0.05. AU: arbitrary units

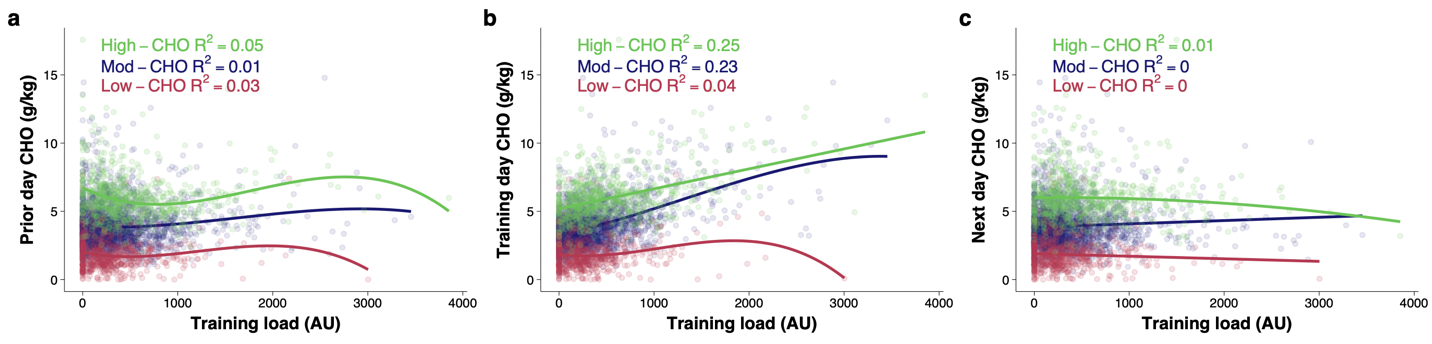

**Supplemental Fig. 7** Daily carbohydrate (CHO) intake relative to training load for the day before (a), day of (b) and day following (c) a given training load (calculated as the product of session rating of perceived exertion (sRPE) and exercise duration in minutes) divided by 10, separated by habitual diet. Best-fit regression lines based on univariable linear mixed effects models are shown for each diet group, with fit indicated as marginal R^2^. AU: arbitrary units
